# Patterns of maternal transport in a state with levels of maternal care and no formal perinatal regions

**DOI:** 10.64898/2026.04.20.26351263

**Authors:** Jingyu Li, Lauren N. Steimle, Margaret Carrel, Riley A. Byrd, Stephanie M. Radke

**Author notes:** **Corresponding Author** Lauren N. Steimle, PhD, Address: 755 Ferst Dr NW, Atlanta, GA 30318. **Declarations Ethics Approval** IRB approval was obtained from the Georgia Institute of Technology (Protocol H23091). **Data Availability** We are unable to share the data due to our data use agreement. **Conflict of Interest** The authors report no conflict of interest. **Authors’ Contributions** Jingyu Li: conceptualization, data curation, methodology, formal analysis, validation, writing – original draft, visualization. Lauren N. Steimle: data acquisition, conceptualization, methodology, validation, writing – original draft, project administration, funding acquisition, supervision. Margaret Carrel: conceptualization, validation, writing – review and editing. Riley Byrd: validation, writing – review and editing. Stephanie M. Radke: conceptualization, validation, writing – review and editing.

## Abstract

**Purpose:** To characterize maternal transport patterns in Iowa, a state with levels of maternal care and without formal perinatal regions, and assess whether transport decisions reflect efficient, risk-appropriate coordination.

**Methods:** We analyzed 2010-2023 Iowa birth records, which included 2,251 maternal transports between obstetric facilities across 106 unique routes. We characterized transport patterns and applied a community detection algorithm to identify “communities” of obstetric facilities that disproportionately transport among themselves.

**Findings:** Suburban and rural counties have elevated transport rates compared to urban counties. 2,189 transports (97%) were from lower-to higher-level facilities. Among these, 2,037 (93%) were to Level III tertiary care centers. 567 transports (25.2%) bypassed a closer facility offering an equivalent or higher level of care than its destination facility. Health system affiliation was associated with bypassing transport, indicating potential organizational rather than purely geographic drivers of transport decisions. Three “communities” of obstetric facilities largely shaped by geographic proximity were identified.

**Conclusions:** Although Iowa does not have formal perinatal regions, patterns of maternal transport are mostly in line with three *de facto* regions. Some potential inefficiencies were identified, such as obstetric facilities transporting to a farther facility when a closer facility offered the same level of care or higher. These findings may help identify opportunities to enhance care coordination among obstetric facilities, optimize maternal transport networks, and improve regionalization of maternal care.

## Introduction

The United States (U.S.) has the highest rates of maternal mortality in high-income countries.^1^ However, as many as 80% of pregnancy-related deaths are deemed preventable.^2^ Among those, more than 20% of preventable deaths are attributed to system-level factors such as gaps in care coordination and the absence of policies and procedures.^3^ This has resulted in calls for systems-level strategies to more effectively deliver maternity care.^4^

Perinatal regionalization, or “risk-appropriate care,” has been a widely recognized strategy for improving maternal and neonatal outcomes. Under this strategy, obstetric facilities are designated by *levels of care* according to their available resources and equipment suitable for handling different maternal comorbidities and pregnancy complications. Regionalization encourages the coordination of care across a geographic area by referring the complex cases to higher-level facilities while ensuring low-risk pregnant people have local options for their birthing facility.^5^

Since 1970s, the March of Dimes Committee on Perinatal Health has outlined a model for regionalized perinatal care.^6^ However, perinatal regionalization has historically focused on neonatal care, and formal maternal regionalization systems are only recently emerging. The first formal guidelines for maternal levels of care were published by the American College of Obstetricians (ACOG) and the Society for Maternal-Fetal Medicine (SMFM) in 2015 and were then reaffirmed in 2019.^7,8^ Recent studies have discussed the promise of maternal regionalization systems, including improved access to risk-appropriate care, reductions in preventable maternal morbidities, and more efficient use of limited high-level resources.^9–11^ States are in the process of developing or implementing levels of maternal care, and as of September 2025, 17 of them had established levels of maternal care guidelines.^12^

Maternal transport is a critical component of regionalized perinatal care. Effective interfacility transport ensures that patients are transferred to facilities with appropriate levels of care for their needs in a timely manner. As of 2019, 37 states had established maternal transport policies.^13^ While some states (such as Georgia and California) have formal perinatal regions within which transports are coordinated, others lack those, resulting in decentralized transport systems.^14–16^ While there has been evidence that providers face challenges in decentralized transport systems, it is not known if these decentralized systems also have broader implications for inefficient use of healthcare resources.^17^

In this study, we characterized maternal transport patterns in Iowa, a state with designated perinatal levels of care and no formal perinatal regions. Iowa has had voluntary designation of levels of perinatal care since 1973.^18^ In 2025, this system was updated to create separate levels of maternal and neonatal care, from Level I to Level IV, aligning with ACOG guidelines.^19^ During the study period, Iowa obstetric facilities were designated under the historical three-tiered system of Perinatal Levels of Care from Level I to Level III, the highest level of care under the state’s Perinatal Levels of Care system at the time.^20^ While Iowa’s facilities are designated with a level of care, the state does not have formally defined perinatal regions to guide care coordination between facilities. We aimed to reveal patterns of maternal transport and identify clusters of facilities that operate as *de facto* regions using a network analysis of maternal transport.^15^ By understanding to what extent the maternal care systems have self-organized and where inefficiencies could exist, our study points to opportunities to design more efficient regionalized systems of maternal care.

## Materials and Methods

### Data

We analyzed Iowa birth records from January 1^st^, 2010, to December 31^st^, 2023. Unique deliveries were identified by collapsing records with the same maternal residential address and infant date of birth. Maternal transports were identified by excluding deliveries with unknown transport status, missing origin or destination facility, or involving non-obstetric locations. Detailed inclusion criteria are shown in **Figure 1**.

**Figure 1.**
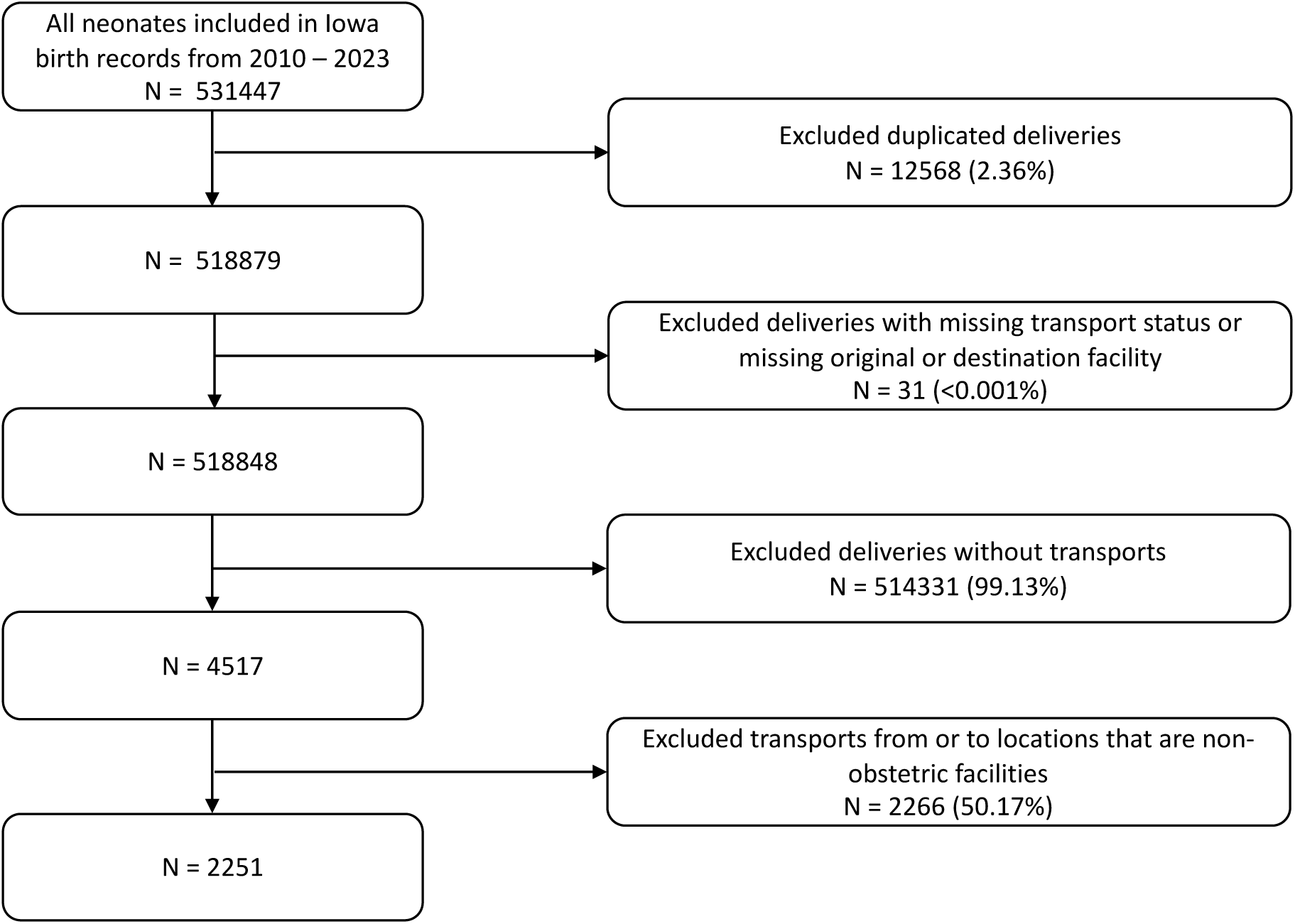
Data inclusion criteria of Iowa birth records, 2010-2023.

### Patient characteristics associated with maternal transport

We divided the study population (n = 518,848) into two groups based on whether maternal transport occurred prior to delivery or not. We compared maternal demographic characteristics, clinical risk factors, and delivery-related conditions between transport and non-transport deliveries. Rural/urban designation of maternal residence used in this study was classified at the county level using 2013 Rural–Urban Continuum Codes (RUCCs): urban (RUCC 1–3), suburban (RUCC 4–6), or rural (RUCC 7–9). We used logistic regression to assess associations between patient characteristics and maternal transport.

### Characterizing transport patterns

We examined variation in transport rates across the state and characterized transport patterns by levels of care and destination facility.

### Rates of transport by county

To examine variation in the transport rates across the state, we reported transport rates per 1,000 county resident births averaged over urban, suburban, and rural counties.

### Rates of transport among different levels of care

We characterized transport patterns among facilities offering each level of care. For the following analyses, we focused only on maternal transports among obstetric facilities during the study period. We classified each facility according to its designated perinatal level of care of each facility under Iowa’s historical three-tiered system of Perinatal Levels of Care (Level I-III).^20^ For each transport, we recorded the maternal level of care of both the origin and destination facilities. Then, we categorized each transport as an upward transport (from a lower-to higher-level facility), a lateral transport (between facilities of the same level), or a back transport (from a higher-to lower-level facility).

### Potential reduction in transport distance with a Level I to Level II transport policy

Several states recommend coordinating maternal transports from a lower-level facility to a Level III-or-higher facility.^14,21,22^ However, these guidelines may result in some patients who could be appropriately managed at a Level II facility being transported farther to receive care at a Level III facility. This is a potential missed opportunity within a regionalized system and contradicts the value of having patients receive risk-appropriate care as close to home as possible. To consider the potential benefits of adopting a Level I to Level II transport policy, we partitioned transports from Level I to Level III into two cases: (1) deliveries requiring Level III care (gestational age < 34 weeks); (2) those potentially appropriate for Level II care (gestational age ≥ 34 weeks).^20^ We further partitioned the latter into: (2a) cases where the destination Level III facility was the closest Level II-or-higher option, and (2b) cases where a closer Level II facility was available. For cases in (2b), we determined the reduction in transport distance if patients had been transported to the closest Level II facility rather than a Level III facility. We estimated the driving distance between obstetric facilities using Google Maps Platform’s Distance Matrix API. This analysis provides a best-case estimate of distance reductions under a policy supporting Level I to Level II transports.

### Characterizing transports that bypass a closer risk-appropriate facility

In some cases, patients were transported to a destination facility that was farther than another facility offering the same level or higher level of care (hereafter, a *bypassing* transport). We characterized the frequency at which bypassing transports occurred. Because the clinical risk level of each transported patient was not available in the birth records, we used the destination facility’s maternal level of care as a proxy for the level of care required. Under this assumption, any facility closer to the patient’s origin that provided at least the destination facility’s level of care was considered to be *risk-appropriate*.

Recognizing that it may be reasonable for facilities to opt to transport a patient slightly farther than the closest risk-appropriate facility, we performed a sensitivity analysis that varied the threshold upon which bypassing is defined. That is, we defined a bypassing threshold of 𝑥 miles and stated that a transport bypassed its closest risk-appropriate facility if it transported its patient to a facility farther than 𝑥 miles beyond the closest risk-appropriate facility. We then varied the bypassing threshold 𝑥.

To identify factors associated with bypassing transports, we fitted multivariable logistic regression models with bypassing as the binary outcome variable. Covariates included patients’ insurance status, distance between origin and destination facilities, and health system affiliation of the origin and destination facilities, which was obtained through online public reports and then manually verified by one domain knowledge expert. We did not consider patient characteristics as transport decisions are primarily made at the facility level rather than at the patient level.

To further measure the influence of health system affiliation on bypassing transports, we repeatedly fitted logistic regression models in which the independent variable was whether the origin and destination facilities belonged to the same health system. For each model, the dependent variable was bypassing status defined under different values of the distance thresholds (in miles) used to define bypassing, 𝑥. This analysis evaluated whether facilities were more likely to transport patients farther when the origin and destination facilities were within the same health system.

### Characterizing facilities with fragmented transport patterns

We define a facility to have a fragmented transport pattern if it routinely transports patients to more than one destination facility with the same level of care. We quantified this behavior by calculating the share of transports directed to the most frequently used destination facility for each origin–destination level-of-care pair. Facilities were classified as having fragmented transport patterns if the dominant destination facility accounted for less than 80% of transports. This threshold was chosen to distinguish between systematic transport to multiple destination facilities and occasional deviations.

### Identifying *de facto* maternal transport regions via community detection

Given that Iowa does not have formal perinatal regions, we sought to identify whether any “informal” regions are observed in the state’s patterns of maternal transport. We used a network-based community detection method to identify empirical (hereafter, “*de facto*”) maternal transport regions. These *de facto* regions are groups of facilities that coordinate maternal transports more frequently amongst themselves than with other hospitals in the network. We followed the same procedures to construct the maternal transport network and identify communities, as described in Li et al., 2025.^15^

To detect clusters of facilities that interact more frequently with one another than with the rest of the network, we applied the Louvain algorithm, which iteratively partitions the network into groups by maximizing modularity, a measure of the density of connections within communities relative to between communities.^23,24^ Facilities grouped within the same community were interpreted as belonging to the same *de facto* maternal transport region. The resulting de facto maternal transport regions reflect empirically observed maternal transport patterns in practice. For this study, we let the algorithm decide the optimal number of regions based on the modularity of the derived communities using directed maternal transport network graphs.

We quantified the extent to which maternal transports occurred within and across these empirically derived *de facto* regions. Each transport was assigned to one of two categories based on the detected community membership of the origin and destination facilities. “Intra-region” transports were defined as transports in which both facilities belonged to the same detected *de facto* region, whereas “inter-region” transports were defined as transports in which the origin and destination facilities belonged to different detected regions. To characterize these patterns, we calculated the proportion and volume of transports falling into each category.

Multivariate logistic regression model was built using R (version 4.5.2). All other analyses presented were performed in Python (version 3.11.14). Community detection and the visualization were implemented using NetworkX package in Python.^25^ We used a significant threshold of 0.05 throughout this study.

## Results

### Data

The study included 518,848 unique deliveries, 2,251 (0.43%) of which involved a maternal transport between obstetric facilities (**Figure 1**). Among all included deliveries (mean [SD] maternal age: 28.3 [5.45] years), 85% were to White people, 6.3% to Black or African-American people, 3.5% to Asian people, 0.8% to American Indian or Alaska Native. 3.6% of these deliveries had a breech presentation, and 29.2% were Cesarean deliveries (**Table 1**).

**Table 1.** Patient characteristics in the study population.

|  | Non-transport<br>(n = 514,331) | Transport<br>(n = 4,517) | Total<br>(n = 518,848) | Unadjusted OR<br>(95% CI) | p value |
| --- | --- | --- | --- | --- | --- |
| **Age in years, mean (SD)** | 28.3 (5.44) | 27.7 (6.08) | 28.3 (5.45) | 0.980(0.975-0.986) | <0.001 |
| **Race, n (%)** |  |  |  |  |  |
| White | 437,164 (85%) | 3,704 (82%) | 440,868 (85%) | ref |  |
| Black | 32,457 (6.3%) | 422 (9.3%) | 32,879 (6.3%) | 1.535 (1.387-1.698) | <0.001 |
| American Indian/Alaska Native | 4,178 (0.8%) | 73 (1.6%) | 4,251 (0.8%) | 2.062 (1.633-2.605) | <0.001 |
| Asian or Pacific Islander | 17,804 (3.5%) | 141 (3.1%) | 17,945 (3.5%) | 0.935 (0.790-1.107) | 0.433 |
| **Ethnicity, n (%)** |  |  |  |  |  |
| Non-Hispanic | 463,184 (90.1%) | 4,089 (90.5%) | 467,273 (90.1%) | ref |  |
| Hispanic | 51,097 (9.9%) | 428 (9.5%) | 51,525 (9.9%) | 0.949 (0.859-1.049) | 0.303 |
| **Urbanicity of mother's residence<sup>a</sup>, n (%)** |  |  |  |  |  |
| Urban | 321,643 (62.5%) | 1,673 (37.0%) | 323,316 (62.3%) | ref |  |
| Suburban | 114,458 (22.3%) | 1813 (40.1%) | 116,271 (22.4%) | 3.045 (2.849-3.256) | <0.001 |
| Rural | 78,082 (15.2%) | 1,029 (22.8%) | 79,111 (15.2%) | 2.534 (2.343-2.739) | <0.001 |
| **Education Level, n (%)** |  |  |  |  |  |
| 8<sup>th</sup> Grade or Less | 16,643 (3.2%) | 127 (2.8%) | 16,770 (3.2%) | ref |  |
| 9<sup>th</sup> through 12<sup>th</sup> Grade; No Diploma | 43,800 (8.5%) | 605 (13.4%) | 44,405 (8.6%) | 1.810 (1.494-2.194) | <0.001 |
| High School or GED | 116,829 (22.7%) | 1,352 (29.9%) | 118,181 (22.8%) | 1.517 (1.263-1.820) | <0.001 |
| College but No Degree | 94,481 (18.4%) | 890 (19.7%) | 95,371 (18.4%) | 1.234 (1.024-1.488) | 0.027 |
| Associate | 65,746 (12.8%) | 547 (12.1%) | 66,293 (12.8%) | 1.090 (0.898-1.323) | 0.382 |
| Bachelor | 122,807 (23.9%) | 723 (16.0%) | 123,530 (23.8%) | 0.772 (0.638-0.932) | 0.007 |
| Master | 39,693 (7.7%) | 192 (4.3%) | 39,885 (7.7%) | 0.634 (0.506-0.794) | <0.001 |
| Doctorate | 13,646 (2.7%) | 67 (1.5%) | 13,713 (2.6%) | 0.643 (0.478-0.866) | 0.004 |
| **Payor<sup>b</sup>, n (%)** |  |  |  |  |  |
| Medicaid | 198,514 (38.6%) | 2,321 (51.4%) | 200,835 (38.7%) | ref |  |
| Private | 292,769 (56.9%) | 1,967 (43.5%) | 294,736 (56.8%) | 0.575 (0.541-0.610) | <0.001 |
| Self-pay | 15,746 (3.1%) | 121 (2.7%) | 15,867 (3.1%) | 0.657 (0.547-0.790) | <0.001 |
| Others | 7,212 (1.4%) | 106 (2.3%) | 7,318 (1.4%) | 1.257 (1.033-1.529) | 0.022 |
| **Tobacco use, n (%)** |  |  |  |  |  |
| No | 429,524 (83.5%) | 3,414 (75.6%) | 432,938 (83.4%) | ref |  |
| Yes | 84,807 (16.5%) | 1,103 (24.4%) | 85,910 (16.6%) | 1.636 (1.528-1.752) | <0.001 |
| **Number of Medical Risk Factors<sup>c</sup>, mean (SD)** | 0.374 (0.619) | 0.772 (0.848) | 0.377 (0.622) | 2.014 (1.949-2.082) | <0.001 |
| **Gestational Age<sup>d</sup>, n (%)** |  |  |  |  |  |
| >= 37 weeks | 475,058 (92.4%) | 885 (19.6%) | 475,943 (91.7%) | ref |  |
| < 37 weeks | 38,781 (7.5%) | 3,629 (80.3%) | 42,410 (8.2%) | 50.231(46.639-54.1) | <0.001 |
| **Plurality, n (%)** |  |  |  |  |  |
| 1 | 506,060 (98.4%) | 4,071 (90.1%) | 510,131 (98.3%) | ref |  |
| >=2 | 8,271 (1.6%) | 446 (9.9%) | 8,717 (1.7%) | 6.703 (6.064-7.409) | <0.001 |
| **Fetal Presentation, n (%)** |  |  |  |  |  |
| Cephalic | 493,680 (96%) | 3,705 (82.0%) | 497,385 (95.9%) | ref |  |
| Breech | 18,069 (3.5%) | 755 (16.7%) | 18,824 (3.6%) | 5.568 (5.141-6.029) | <0.001 |
| Other | 2,497 (0.5%) | 56 (1.2%) | 2,553 (0.5%) | 2.988 (2.289-3.902) | <0.001 |
| **Final Method of Delivery, n (%)** |  |  |  |  |  |
| Vaginal | 365,096 (71.0%) | 2,217 (49.1%) | 36,7313 (70.8%) | ref |  |
| Cesarean | 149,220 (29.0%) | 2,300 (50.9%) | 151,520 (29.2%) | 2.538 (2.394-2.692) | <0.001 |
| **Infant Transfer after Birth, n (%)** |  |  |  |  |  |
| No | 506,756 (98.5%) | 4,412 (97.7%) | 511,168 (98.5%) | ref |  |
| Yes | 7,570 (1.5%) | 105 (2.3%) | 7,675 (1.5%) | 1.593 (1.311-1.936) | <0.001 |
| **Abnormal Condition relating to NICU Admission, n (%)** |  |  |  |  |  |
| No | 473,420 (92.0%) | 1102 (24.4%) | 474,522 (91.5%) | ref |  |
| Yes | 40,888 (7.9%) | 3,415 (75.6%) | 44,303 (8.5%) | 35.881 (3.350-3.843) | <0.001 |

Abbreviations: OR, odds ratio; CI, confidence interval; SD, standard deviation; ref, reference level.
\* Significant at p=0.05; \*\*: significant at p=0.01; \*\*\*: significant at p=0.001
† Patients with missing information were not reported in the table.
- Urbanicity of mother's residence: we categorized mother's residence by linking census block group to the Rural-Urban Continuum Codes (RUCCs): urban (RUCCs of 1-3), suburban (RUCCs of 4-6), and rural (RUCCs of 7-9). - Other payor includes Indian Health Service, CHAMPUS/TRICARE, Other Government (federal, state, local), or OB indigent program/Other. - Medical risk factors: pre-pregnancy diabetes, gestational diabetes, eclampsia, pre-pregnancy hypertension, gestational hypertension, previous preterm birth, and previous Cesarean delivery. - Preterm birth refers to any deliveries with gestational age less than 37 weeks.

### Patient characteristics associated with maternal transport

**Table 1** summarizes important maternal and neonatal characteristics for non-transport and transport deliveries. Multiple factors are significantly associated with transports (p < 0.05). Compared with non-transport births, transported patients differed across several demographic factors. The transport group had higher proportions of Black (9.3% in transport vs. 6.3% in non-transport) and American Indian/Alaska Native patients (1.6% in transport vs. 0.8% in non-transport), and a lower proportion of urban residents (37% in transport vs. 62.5% in non-transport). Transported patients were more often Medicaid beneficiaries (51.4% in transport vs. 38.6% in non-transport). Ethnicity (Hispanic or Non-Hispanic) was not significant between transport and non-transport groups.

Clinical characteristics also differed significantly between the transport and non-transport groups. Transported patients had higher rates of tobacco use during pregnancy (24.4% in transport vs. 16.5% in non-transport) and a greater mean number of documented medical risk factors (0.77 in transport vs. 0.37 in non-transport). Transports included a significantly larger share of preterm births (80.3% in transport vs. 7.5% in non-transport), multifetal pregnancies (9.9% in transport vs. 1.6% in non-transport), and breech fetal presentations (16.7% in transport vs. 3.5% in non-transport). Cesarean deliveries were more common among transported patients (50.9% in transport vs. 29% in non-transport), and they had higher rates of infant transfer after birth (2.3% in transport vs. 1.5% in non-transport) and NICU admission due to abnormal neonatal conditions (75.6% in transport vs. 7.9% in non-transport).

### Characterizing of transport patterns

#### Rates of maternal transport by county

County-level maternal transport rates averaged 11.50 [SD: 7.32] per 1,000 county resident births and were highest in suburban counties (13.43 [SD: 7.59]), followed by rural counties 12.68 [SD: 7.09], with substantially lower rates in urban counties 5.92 [SD: 4.17] (**Figure 2**).

**Figure 2.**
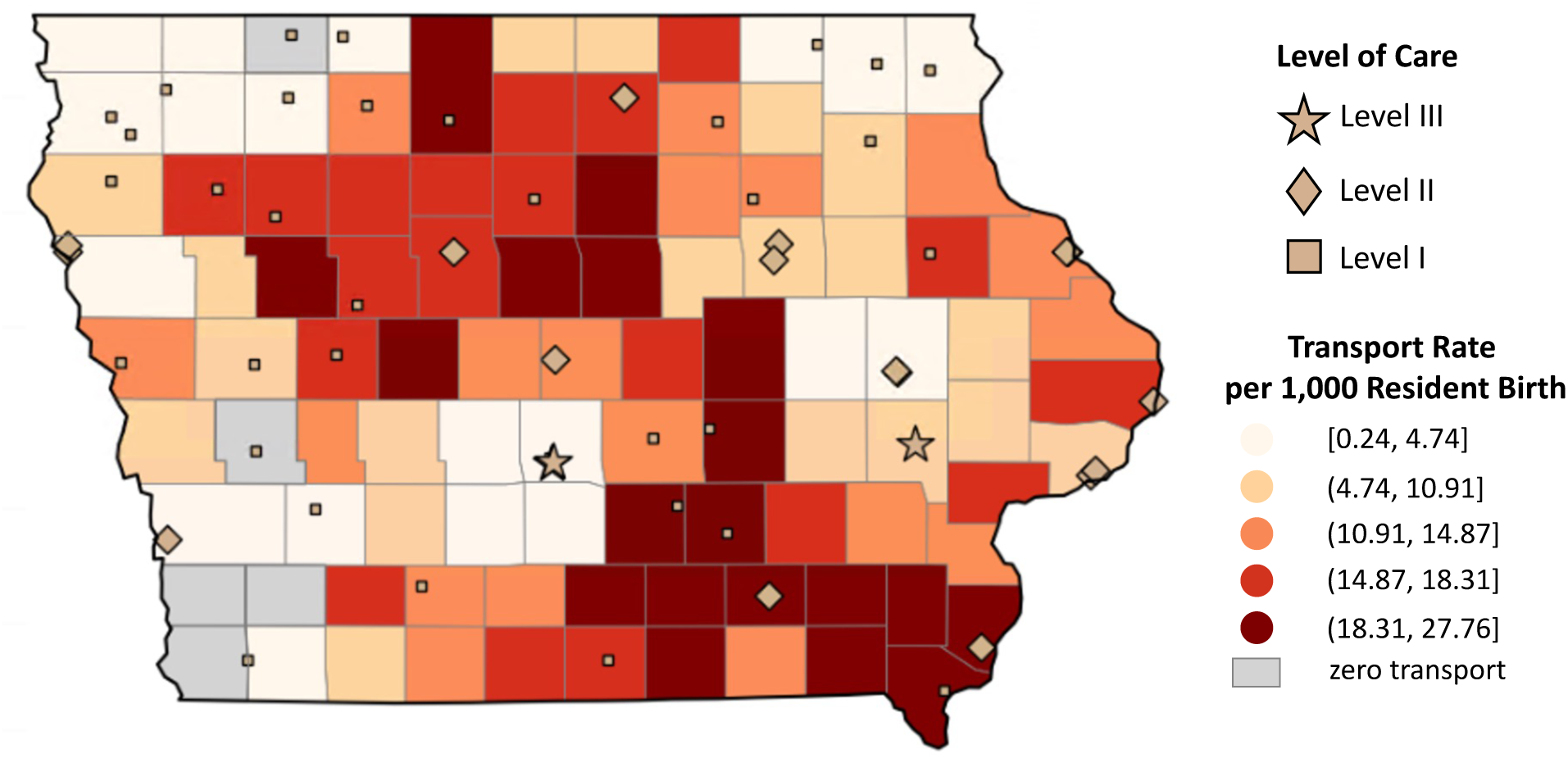
County-level maternal transport rates per 1,000 county resident births in the state of Iowa. The average transport rate across all counties in Iowa was 11.50 [SD: 7.32] per 1,000 resident births. Counties are shaded by quintile of transport rate, with darker shading indicating higher rates; counties with zero maternal transports are shown in grey. Obstetric facilities are indicated by square (Level I), diamond (Level II), and star (Level III) symbols under the state’s historical three-tiered system of Perinatal Levels of Care during the study period.

#### Rates of transport among different levels of care

Among included transports between obstetric facilities, 2,189 (97%) were from lower-level facilities to higher-level facilities, with 2,037 (93%) transports directed to Level III facilities (**Table 2**).

**Table 2.** Maternal transport by levels of care.

| **Origin Level of Care** | **Destination Level of Care** |  |  |
| --- | --- | --- | --- |
|  | Level I | Level II | Level III |
| Level I | 4 (0.4%) | 152 (16.7%) | 754 (82.9%) |
| Level II | 5 (0.4%) | 27 (2.0%) | 1283 (97.6%) |
| Level III | 11 (15.5%) | 2 (2.8%) | 58 (81.7%) |

#### Potential reduction in transport distance with a Level I to Level II transport policy

Among 754 transports from Level I to Level III facilities, 307 (40.7%) transports had gestational age less than 34 weeks. For those transports involving a patient with a gestational age of at least 34 weeks, 261 (58.4%) bypassed a closer Level II facility, and the median incremental distance to bypass for a Level III is 49.7 miles (25^th^ percentile: 30.6 miles, 75^th^ percentile: 59.6 miles).

#### Characterizing transports that bypass a closer risk-appropriate facility

567 (25.2%) transports along 66 unique transport routes bypassed a closer facility with the same or higher level of care. When restricting to routes with at least 5 patients, 487 transports (21.6%) along 23 unique routes bypassed a closer facility of the same level of care or higher. Among these, 358 (73.5%) along 15 unique routes occurred when the bypassed facility was within 15 miles of the origin (**Figure 3**).

**Figure 3.**
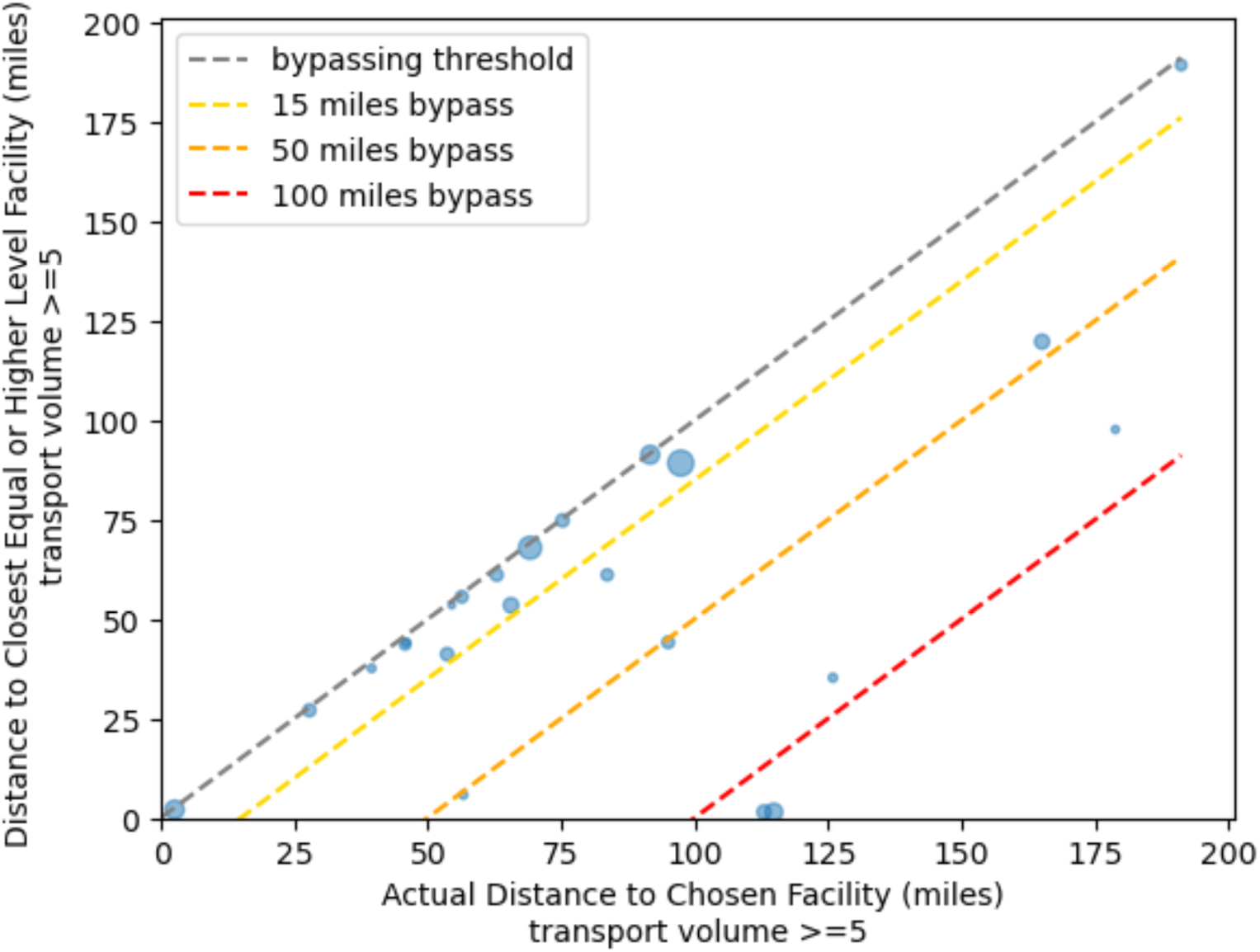
Comparison of actual transport distance and distance to closest facility offering the same or higher level of care. Each circle represents a unique route, with circle size proportional to the number of transports along that route. Dashed diagonal lines indicate bypass thresholds at incremental distances: gray (no bypass; actual transport was to the closest facility of that level or higher), yellow (15 miles), orange (50 miles), and red (100 miles). Routes below the gray line indicate bypassing of a closer facility with the same or higher level of care. Transport routes with fewer than five patients are masked.

As shown in **Table 3**, bypassing transports were more likely to occur when the origin and destination facilities belong to the same health system (OR: 3.281, 95 CI%: 2.541-4.238, p < 0.001). Conversely, bypassing transport was less likely to occur when the newborn had abnormal conditions relating to Neonatal Intensive Care Unit (NICU) admission compared to those who do not have abnormal condition relating to NICU admission (OR: 0.730, 95 CI%: 0.581-0.917, p = 0.007).

**Table 3.** Factors associated with bypassing transport.

| **Variable** | **Adjusted Results<sup>a</sup>** |  | **Unadjusted Results<sup>b</sup>** |  |
| --- | --- | --- | --- | --- |
|  | **OR (95% CI)** | **p value** | **OR (95% CI)** | **p value** |
| **Payor (ref: Mediciad)** |  |  |  |  |
| Private | 0.945 (0.774-1.153) | 0.578 | 0.925 (0.762-1.124) | 0.435 |
| Self-pay | 1.507 (0.827-2.745) | 0.180 | 1.633 (0.914-2.918) | 0.980 |
| Others | 0.277 (0.108-0.708) | 0.007** | 0.278 (0.110-0.703) | 0.007 |
| **Abnormal Condition relating to NICU Admission (ref: No)** |  |  |  |  |
| Yes | 0.730 (0.581-0.917) | 0.007** | 0.727 (0.582-0.908) | 0.005 |
| **Transport between facilities within the same health system (ref: No)** |  |  |  |  |
| Yes | 3.281 (2.541-4.238) | <0.001 *** | 3.297 (2.557-4.251) | <0.001 |

Abbreviations: OR Odds ratio, CI Confidence interval, ref reference level, NICU Neonatal intensive care unit
a. p values were derived from the multivariable logistic regression model, adjusting for all covariates listed in the table
b. p values were obtained from univariate logistic regression models

Figure 4 illustrates how the effect of health system affiliation on bypassing varies by incremental travel distance threshold 𝑥 (in miles). Facilities were more likely to bypass closer options for a destination facility within the same system when the additional distance was reasonably small. For example, facilities were 3.3 times more likely to bypass a facility with the same or higher level of care in favor of a farther facility within the same health system at 𝑥 = 0 (p < 0.001). As the incremental distance threshold increased, the magnitude of this effect declined, indicating a diminishing willingness to transport patients farther to keep them in the same health system. Facilities were only 1.6 times more likely to bypass a closer facility when a same-system destination was at least 12 miles farther away (p = 0.008). Beyond 13 miles, the odds ratios of bypassing transport for a farther facility within the same health system approached 1 and were no longer statistically significant.

**Figure 4.**
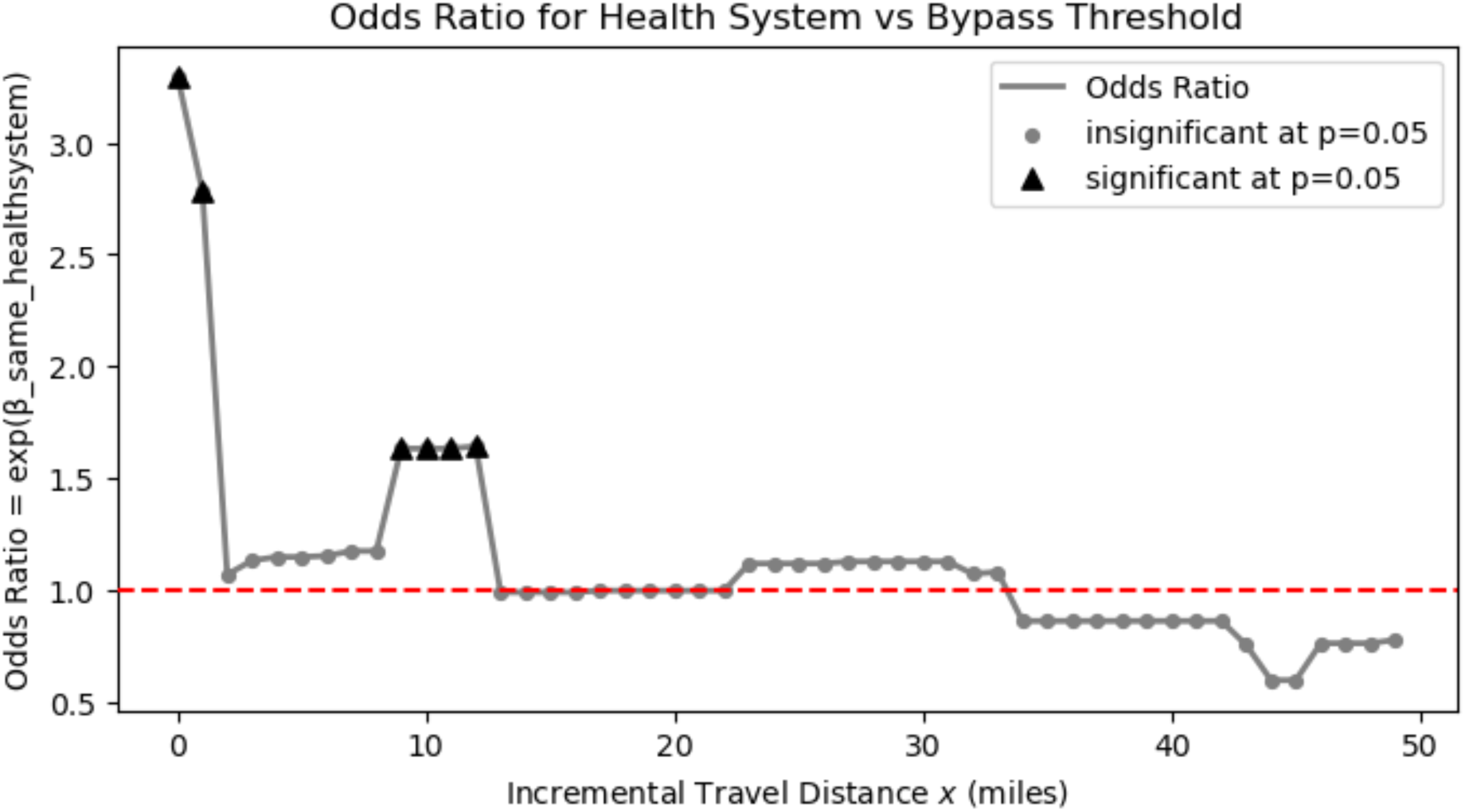
Odds ratio for transport within the same health system under incremental travel distance for bypassing (0-50 miles). Grey circles and black triangles represent odds ratio insignificant and significant at p value of 0.05. The dashed horizontal line in red at an odds ratio of 1.0 represents no association.

#### Characterizing facilities with fragmented transport patterns

**Table 4** details the facility transport profile for upward transports and lateral transport between Level III facilities. Although a few facilities transported to at least two facilities with the same level of care, facilities routinely transporting along multiple routes were rare. The percentage of origin facilities with fragmented transport patterns was the highest in Level I to Level III transports (22.7%), followed by Level I to Level II (15.4%) and Level II to Level III transports (14.3%).

**Table 4.** Fragmented transport patterns among origin facilities by level of care.

| Transport direction | Number of origin facility | Facility with multiple routes<sup>a</sup> (n%) | Facility with dominant route share < 0.8<sup>b</sup> (n%) | Average number of routes with share between 0.2 and 0.8<sup>c</sup> |
| --- | --- | --- | --- | --- |
| Level I to Level II | 13 | 4 (30.8) | 2 (15.4) | 2 |
| Level I to Level III | 22 | 11 (50) | 5 (22.7) | 2 |
| Level II to Level III | 14 | 5 (35.7) | 2 (14.3) | 2 |
| Level III to Level III | 2 | 2 (100) | 0 (0) | NA |

a. Number and percentage of origin facilities that transported patients to at least two different destination facilities within the same level of care category.
b. Number and percentage of origin facilities where the most frequently used route accounts for less than 80% of transports, indicating multiple routine transport routes to facilities with the same level of care.
c. Among facilities with dominant route share less than 0.8, the average number of unique destination routes with a share between 0.2 and 0.8.

#### Identifying *de facto* maternal transport regions via community detection

Community detection identified three *de facto* regions driven primarily by geographic proximity: West (orange), Central (blue), and East (green) (Figure 5). 2,043 of 2,251 transports (90.8%) occurred within the same *de facto* region, while inter-region transports almost exclusively involved transport to Level III facilities in adjacent regions (**Table 5**).

**Figure 5.**
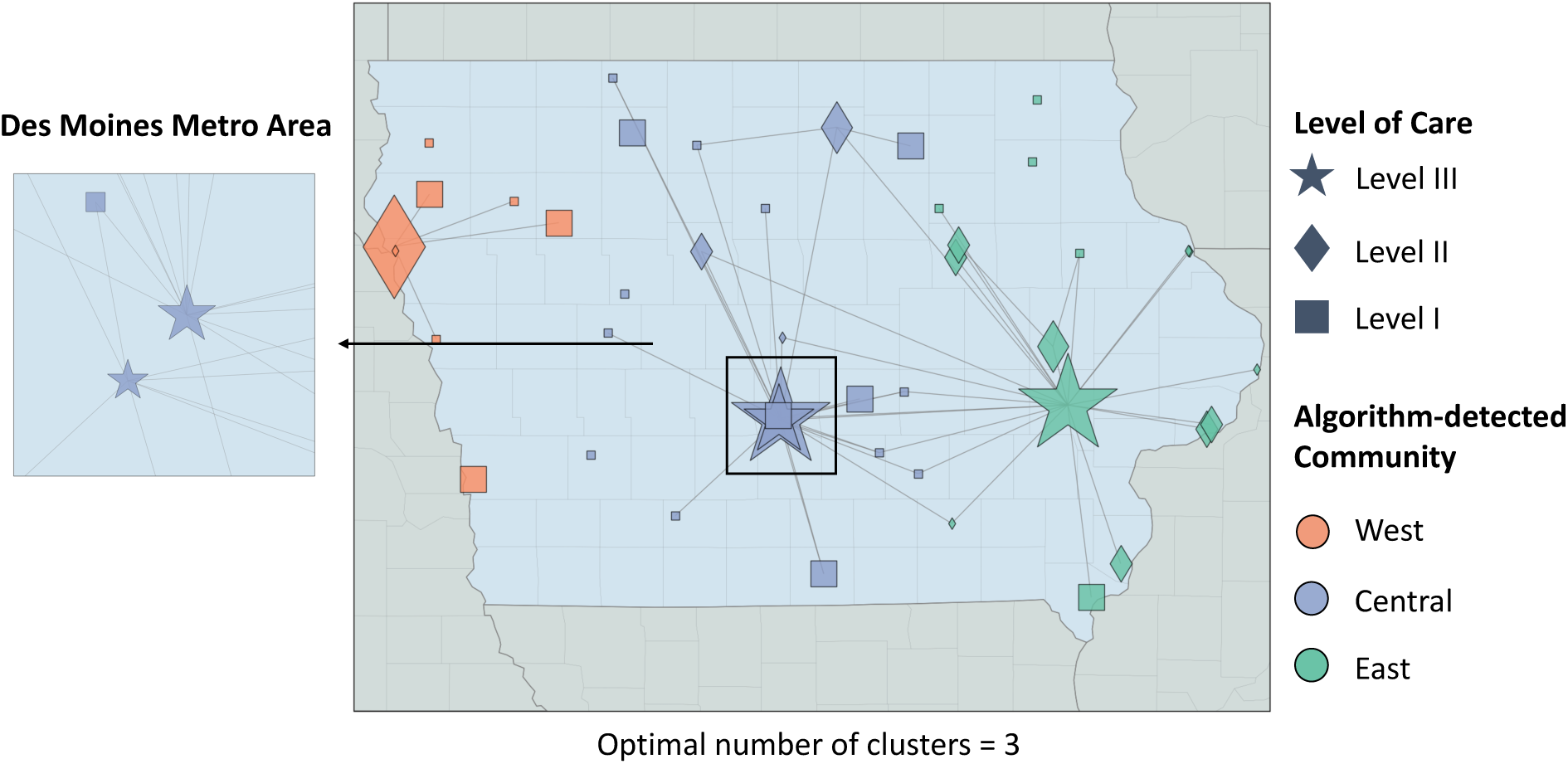
*de facto* regions of maternal transport in Iowa identified using community detection algorithm (optimal number of regions=3, modularity = 0.45). Nodes represent obstetric facilities, with shape indicating designated level of care, size proportional to incoming transport volume, and color indicating their algorithm-detected regions. Edges represent maternal transport between the respective facilities. Direction of the transport is omitted for visual clarity. Transport routes with fewer than five patients are masked.

**Table 5.** Maternal transport by *de facto* regions.

|  | **Destination Region<sup>a</sup>** |  |  |
| --- | --- | --- | --- |
|  | West | Central | East |
| **Origin Region** |  |  |  |
| West | 113 | 1* | 5* |
| Central | 4 | 883 | 162<sup>b</sup> |
| East | 0 | 36<sup>c</sup> | 1047 |

\* Each transport's destination facility was a Level III facility in the corresponding destination region.
a. 201 out of 208 (96.6%) inter-region transport involved transporting to a Level III facility in a different region.
b. 161 out of 162 (99.4%) involved transporting to the Level III facility in the East region.
c. 34 out of 36 (94.4%) involved transporting to the Level III facility in the Central region.

## Discussion

In this study, we characterized maternal transport patterns among obstetric facilities in a predominantly rural state with long-standing perinatal levels of care but no formal perinatal regions. This study adds to the existing literature on maternal transport patterns by examining a state without formal perinatal regions, in contrast to a prior work examining transports in Georgia, a state with formally defined perinatal regions.^15^ This study demonstrated that Iowa’s maternal transport patterns have shown a degree of self-organization into *de facto* regions in the absence of formal regions, but that there may be opportunities to better coordinate maternal transports among obstetric facilities. In particular, we observed that more than 25% of transport bypassed closer, risk-appropriate facilities. Among patients who might only require a Level II facility, more than half bypassed a Level II facility for a farther Level III facility, suggesting possible overutilization of high-level obstetric services. In addition, some facilities transported patients to multiple destination facilities of the same level of care, which could indicate fragmented rather than consistent, coordinated pathways for maternal transport. One possible concern is if these fragmented pathways are results of situations when an origin facility made multiple requests for transfer before identifying a destination facility. However, these patterns may also arise due to patient-level factors, such as preference or specific medical needs. These transports presented potential opportunities to improve regionalized systems of maternal care in Iowa through the strategic design of perinatal regions that better align transport patterns with the principles of delivering risk-appropriate care.

Transport rates varied significantly across the state. Notably, proximity to a Level III facility does not necessarily correspond to lower transport rates. For example, several counties in southeast Iowa exhibited above-average transport rates despite being relatively close to a Level III center. Conversely, some regions located far from any Level III facility did not show elevated transport rates. Several factors could explain this counterintuitive pattern. First, patients who live further away from Level III facilities in Iowa might seek higher levels of care across the state’s borders. For example, in certain regions, such as southwest Iowa, patients may opt to receive subspecialty care at a closer higher-level facility in a neighboring state, such as Nebraska. These interstate transports would not be reflected in our data. Another potential explanation is that a transport requiring long distances may not be an option once complications arise. Thus, our data may be masking instances in which patients are not transported and ultimately do not receive risk-appropriate care.

Maternal transport rates varied by level of rurality across the state. Urban counties had lower transport rates than rural and suburban counties. However, suburban counties exhibited higher transport rates compared to rural counties, which differs from the state of Georgia in which rural counties were shown to have the highest transport rates.^15^ This pattern may reflect the unique position of suburban counties within Iowa’s regionalized systems of maternal care. In Iowa, suburban counties often contain Level I and II obstetric facilities that serve as initial points of care but are geographically proximate to Level III facilities, leading to more frequent interfacility transports as patient risk is reassessed or escalates prior to delivery. In contrast, many rural counties in Iowa are far from the three in-state Level III facilities and might have greater reliance on planned referral for delivery earlier during their pregnancy, which could reduce the observed volume of interfacility transport.

Our study highlights the importance of health systems affiliation in maternal transport decisions. We found that transports were often coordinated among facilities in the same health system, even if there was a closer hospital providing the same desired level of care. The importance of health system affiliation in maternal transport patterns was also observed in Georgia. As discussed in Li et al. 2025, there might be several potential health benefits associated within-system transport, such as the continuity of care provided by shared providers and electronic health records.^15,26^ Additionally, these patterns may also reflect reimbursement policies that incentivize hospitals to keep patients within their health system.^27^ The decision on transport destination must carefully weigh the potential benefits of care coordination against the potential harms, particularly when a much closer risk-appropriate facility outside the system is available. Thus, policymakers should carefully consider how reimbursement policies for maternal transport may incentivize hospitals to transport within their own health system even if a closer risk-appropriate hospital is available.

Our study points to potential opportunities to design more efficient regionalized systems of maternal care, particularly by addressing the potential overutilization of higher-level obstetric facilities. We found that only 40.7% of transports among Level I to Level III facilities had a gestational age of less than 34 weeks that clearly required Level III care. For the remaining cases, more than half bypassed a closer Level II facility. If these same patients were instead referred to their closest Level II facility, half of them would see more than a 49 miles reduction in transport distance. When interpreting these results, we remind the reader that not all of these patients would truly require Level III care. We were not able to obtain other risk factors that would indicate the need for patients to deliver in a Level III facility beyond gestational age. However, the selected cases might provide a look into the potential overutilization of higher-level resources, as a closer Level II facility might have been sufficient to meet clinical needs for some of these patients. Transporting patients from a Level I to a Level III when a Level II is risk-appropriate could present a source of unnecessary increases in travel distance and strain capacity at higher-level facilities.

The extent of fragmented transport patterns varied by the levels of care at origin and destination facilities. These fragmented patterns occurred most frequently from Level I to Level III facilities, among which 22.7% routinely transported patients to more than one destination. While these patterns may reflect clinically informed decision-making in response to patient needs, preferences, or real-time capacity constraints, they may also indicate the lack of formal regions and indicate the absence of stable referral relationships with higher-level facilities. Maternal transport coordination is complex and engages different parties, including the origin provider, destination provider, and transport personnel. This process typically involves checklists across patient conditions and provider capacity.^28,29^ In the absence of formal perinatal regions, destination facilities may vary in their willingness or ability to accept transports due to differences in capacity, clinical thresholds for higher-level care, or perceived obligation to the origin facility.

Transport routes to multiple destination facilities could indicate diffused referral relationships between the original and destination facilities. In this case, destination higher-level facilities may be less likely to engage in shared clinical decision-making or to provide consultation that could facilitate timely acceptance of appropriate transfers. Although our data do not allow us to understand the process by which a destination facility was selected, we point to important future research direction to investigate reasons why certain facilities routinely transport patients to multiple destination facilities with the same level of care. Establishing clear regional affiliations and transport protocols could promote greater accountability, earlier communication, and more consistent transport practices.

Our community-detection algorithm identified three *de facto* regions describing groups of facilities that disproportionately transport amongst themselves. These *de facto* regions are mostly consistent with geographic proximity. By better understanding current patterns of maternal transports, policymakers could design perinatal regions that formalize existing partnerships between obstetric facilities. If new perinatal regions were to be established in Iowa, this analysis suggests that creating a region in the east with a Level II facility as the region center, a region in the middle and in the east with their corresponding Level III facilities as region centers would be in line with the observed transport patterns.

This study has several limitations. First, as a single-state analysis, our results might not generalize to other states with different birthing populations or distribution of obstetric resources. Second, birth records do not contain information on the minimum level of care that was risk-appropriate for the transported patient. As a result, our analyses relied on the level of care at the destination of the transport, which may overestimate the level of care that was clinically appropriate for the patient. For example, a portion of transports greater than 34 weeks may have unmeasured maternal or neonatal factors necessitating Level III care. Future analyses could consider linking birth records and hospital discharge records to better identify the appropriate level of care for each transport and more precisely assess potential overutilization or unnecessary transport distance. Third, we used driving distance instead of driving time when we determined the geographic proximity from the origin facility to the destination facility, which could overestimate transport bypassing. Fourth, our analysis only focused on transports between obstetric facilities. Incorporating maternal transports originating from hospitals without obstetric services or the patient’s home could better capture areas with the most restricted access to care. Fifth, perinatal levels of care were self-reported and assumed to remain constant throughout the study period. This designation might not be aligned to the current national guidelines from ACOG given the time period that was evaluated.^30^ Moreover, facilities may have changed the level of care they were able to provide over the more than 10-year study period. Additionally, we did not explicitly account for geographic clustering within health systems, where facilities may be collocated and affected transport decisions. Finally, our dataset did not describe births that occurred to Iowans in neighboring states, such as South Dakota and Nebraska. Therefore, we could have underestimated transport rates in regions where patients were transported to out-of-state facilities.

In conclusion, our study characterized transport patterns in a state with maternal levels of care but no formal perinatal regions. While Iowa appears to have several *de facto* regions, there are still opportunities to improve the efficiency of the maternal transport system. These results could inform future strategic design of perinatal regions in similar states that do not currently have formal regions.

## Data Availability

We are unable to share the data due to our data use agreement.

